# Developing A Deep Learning Natural Language Processing Algorithm For Automated Reporting Of Adverse Drug Reactions

**DOI:** 10.1101/2021.12.11.21267504

**Authors:** Christopher McMaster, Julia Chan, David FL Liew, Elizabeth Su, Albert G Frauman, Wendy W Chapman, Douglas EV Pires

## Abstract

The detection of adverse drug reactions (ADRs) is critical to our understanding of the safety and risk-benefit profile of medications. With an incidence that has not changed over the last 30 years, ADRs are a significant source of patient morbidity, responsible for 5-10% of acute care hospital admissions worldwide. Spontaneous reporting of ADRs has long been the standard method of reporting, however this approach is known to have high rates of under-reporting, a problem that limits pharmacovigilance efforts. Automated ADR reporting presents an alternative pathway to increase reporting rates, although this may be limited by over-reporting of other drug-related adverse events.

We developed a deep learning natural language processing algorithm to identify ADRs in discharge summaries at a single academic hospital centre. Our model was developed in two stages: first, a pre-trained model (DeBERTa) was further pre-trained on 1.1 million unlabelled clinical documents; secondly, this model was fine-tuned to detect ADR mentions in a corpus of 861 annotated discharge summaries. This model was compared to a version without the pre-training step, and a model finetuned from the ClinicalBERT model, which has demonstrated state-of-the-art performance on other pharmacovigilance tasks. To ensure that our algorithm could differentiate ADRs from other drug-related adverse events, the annotated corpus was enriched for both validated ADR reports and confounding drug-related adverse events using. The final model demonstrated good performance with a ROC-AUC of 0.955 (95% CI 0.946 - 0.963) for the task of identifying discharge summaries containing ADR mentions, significantly outperforming the two comparator models.

## 1. Introduction

Adverse drug reactions (ADRs) are a subset of adverse drug events (ADEs), defined as injuries resulting from drugs administered at therapeutic doses [1]. With an incidence that has not changed over the last 30 years, ADRs are estimated to be responsible for 5-10% of acute care hospital admissions and one of the leading causes of death worldwide [2, 3, 4]. In Australia, ADRs result in approximately 2-4% of hospital admissions and are a significant source of patient morbidity [5].

Due to the challenges of clinical note processing, ADRs are currently reported spontaneously by clinical staff [6] or, less commonly, through the ICD-10 Y40.0-Y59.9 coding for ADEs [7] (see Figure 1). Spontaneous reporting is the predominant way in which ADRs are reported. Whilst it is a relatively accurate method, the under-reporting rate is estimated to range from 82-98% [6]. In Australia, clinician review and reporting of ICD-10 Y40.0-Y59.9 coded ADEs can capture some of these missed ADRs from spontaneous reporting, as ADEs are coded retrospectively by professional clinical coders who scan clinical data to identify patient diagnoses for the purposes of hospital funding. However, using ADE codes for ADR detection is a relatively resource intensive process, with a large volume of ADE events and only a small subset of ADRs. Not all hospitals have the capacity to review all coded records to differentiate the ADRs [8]. Given coding for ADEs is performed by individuals not involved in clinical care, multiple opportunities for inconsistency in ADE reporting arise, including inter-coder variability [9], lack of context, knowledge, and time [2].

**Figure 1:**
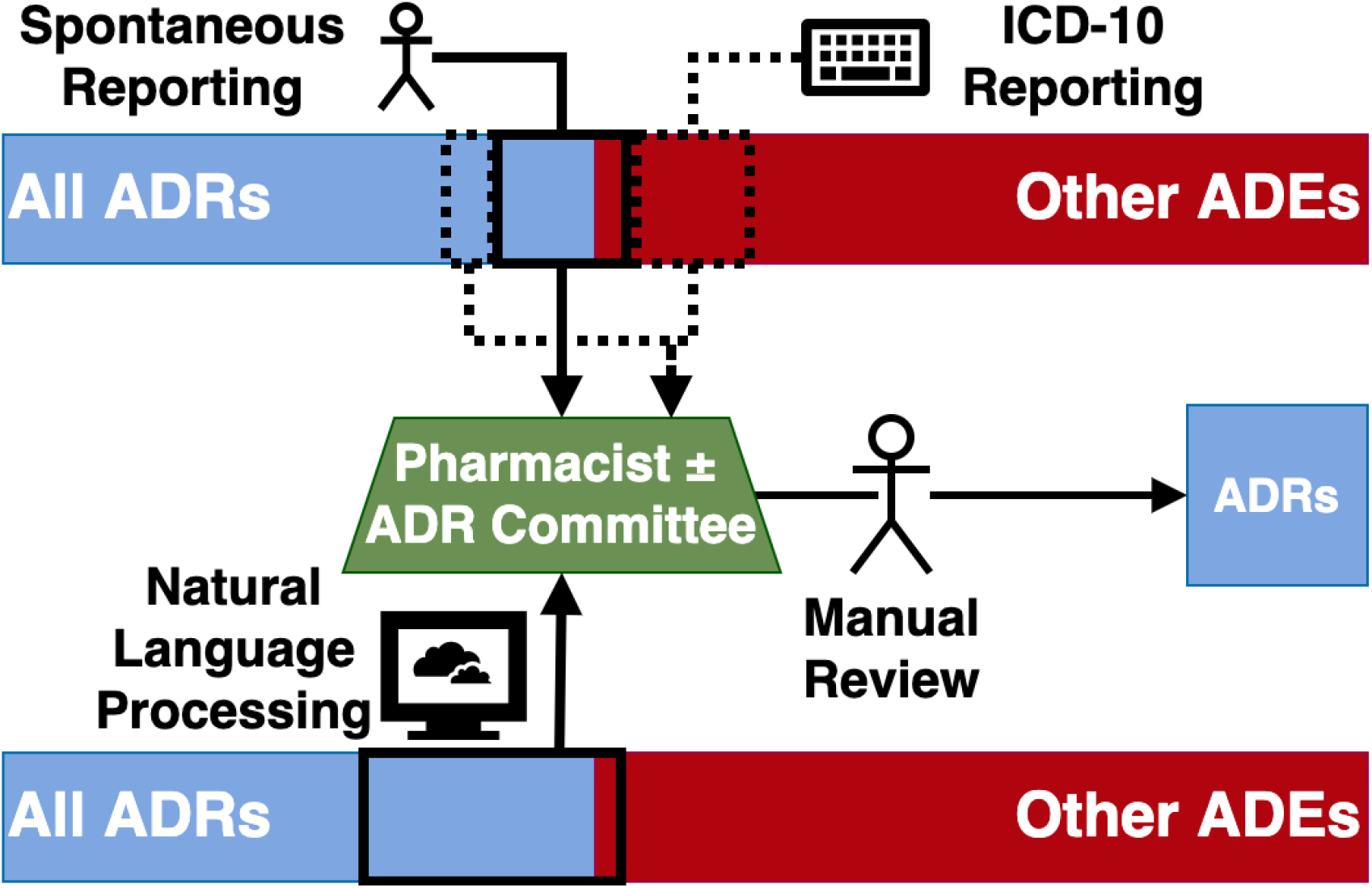
ADR reporting mechanisms. The current state of ADR reporting is primarily based on spontaneous reporting, with previous studies demonstrating a high rate of unreported ADRs. The addition of ICD-10 coding can increase this reporting rate, however many other ADEs are picked up in this process, placing strain on the review process. The aim of an NLP model is to match or exceed the ADR reporting rate from spontaneous reporting plus ICD-10 codes, whilst significantly reducing the burden of other ADE reports.

Although maintaining a comprehensive hospital-based pharmacovigilance program is relatively costly and time-intensive [8], reporting ADRs is an essential component of clinical practice because it contributes to our broader understanding of the safety and risk-benefit profile of medications [2]. Failure to recognise ADRs can lead to delays in changing post-marketing product label warnings and actioning changes [10]. More efficient pharmacovigilance strategies to identify ADRs are required to rival the increasing number of drugs approved through expedited and provisional pathways [11, 12]. For these reasons, there is a need to create new systems to capture ADRs.

Considering that reviewing patient history is the most time-consuming process in maintaining a pharmacovigilance program, automated machine learning models have the potential to be trained to detect ADRs in real-time and flag potential high-risk cases for further review when required [7]. In particular, admissions coded with ADE codes provide a challenging dataset for modle training – manual review of these admissions is tiem-consuming precisely because differentiating ADRs from other ADEs is a difficult task. Furthermore, EMR data is generally inexpensive, accessible, can be obtained without interfering with patient care, and reflects real-world clinical outcomes [2]. In the same vein, machine learning models are currently being explored as methods for predicting ADRs in the preclinical stages of drug development [13].

### 1.1. Similar Work

In recent years there has been increasing interest in using natural language processing (NLP) for the detection of ADEs, a broader group of adverse events that encompases ADRs, as well as poisonings and other medication-related harm. Several datasets have been developed for benchmarking algorithms, most notably the n2c2-2018 ADE [14] and the MADE 1.0 [15] datasets. Four distinct NLP paradigms have been used for ADE detection in these datasets, namely: 1) rule-based, 2) machine learning-based 3) deep learning-based, and 4) contextualized language model-based approaches. The best performance has been seen with deep learning methods [16], in particular using pre-trained large language models (LLMs) such as BERT [17], BioBERT and ClinicalBERT [18].

In contrast, ADR detection has not been as well studied. Distinguishing ADRs from other ADEs is a task that often requires extensive review of clinical notes to exclude both alternative causes and inappropriate medication usage. Attribution of causality hinges on scoring systems like the Naranjo Score [19]. One approach to ADR detection has been to predict the Naranjo Score from the clinical notes [20], however this assumes that the score components have been documented. When these components are not documented, that does not necessarily mean they were absent. This has the potential to introduce a further source of bias from differential documentation. An alternative approach that we propose is to distinguish ADRs from ADEs by using dataset enriched for both, so that our models must learn to differentiate these events in order to attain good performance.

### 1.2. Study Aim

The aim of this study was to develop an NLP algorithm to identify ADRs in discharge summaries and, in particular, to distinguish ADRs from other ADEs to augment ADR reporting (see Figure 1). In doing so, we developed a new corpus of ADR annotations, enriched for both ADRs and ADEs. Like the most successful ADE detection algorithms, we used a deep learning approach, in particular pre-trained LLMs. In order to adapt to the national (Australia) and institutional documentation practices and vocabulary, we further pre-trained our model on a large corpus of unannotated discharge summaries from our institution, prior to fine-tuning on the annotated corpus.

## 2. Methods

### 2.1. Institutional Setting

861 discharge summaries spanning a 5 year period (2015-2020) from the Electronic Medical Record (EMR) were collected and retrieved from the Clinical Research Data Warehouse of a 900-bed metropolitan tertiary teaching hospital network in Melbourne, Australia. All discharge summaries were from admissions coded with a Y40-59 ICD-10 code (ADE code), meaning that all admissions had been assessed by a clinical coder as containing an ADE – including a subset of 231 that had been previously reviewed and validated as true ADRs by our institutional ADR committee.

This model was governed by Austin Health Quality Improvement Number 41519.

### 2.2. Model Training and Evaluation

Figure 2 illustrates the full model training and evaluation process.

**Figure 2:**
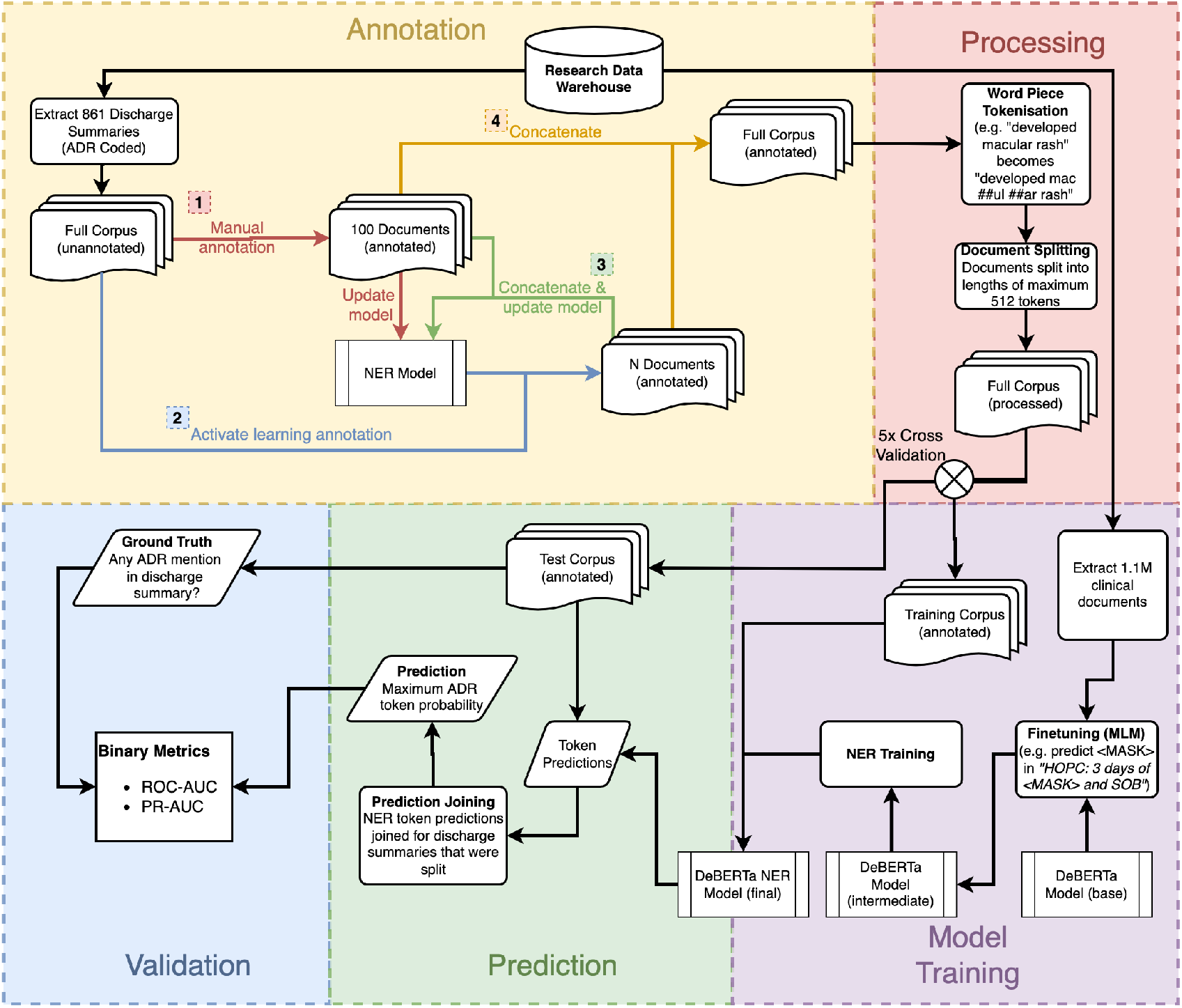
Full NLP Pipeline. Annotation is split into 4 steps performed in order – steps 2 & 3 are repeated in sequence with N discharge summaries each time, until either all data is annotated or the updated NER model shows minimal improvement when more annotated data is added.

#### 2.2.1. Annotation

The Prodigy annotation tool [21] was used to annotate drug names and known adverse drug reactions in the discharge summaries. We achieved rapid annotation of a moderate sized corpus by using active learning, an iterative process whereby annotation is performed in batches. On completion of each batch, an intermediate NLP model was retrained on the current annotations. This model was then used to suggest labels to the annotator, thus focusing the annotator’s attention on the most likely locations of interest within the text. The first set of 100 annotations were performed manually using the ner.manual Prodigy recipe to create an initial annotated corpus. The active learning process was then employed for all future batches (100-200 texts per batch), using the open-source Med7 model [22] as the initial intermediate NLP model. This ner.correct Prodigy recipe was used for all batches using the active learning process. This process was repeated until all documents were annotated. The two labels were ‘DRUG’ and ‘ADR’, following a standardized label scheme (Appendix A) to create Named Entity Recognition (NER) training data for the model. A single annotator was trained to perform all annotations, which were then reviewed by a senior clinician.

#### 2.2.2. Model

We used the DeBERTa model, a LLM neural network architecture that has been shown to outperform BERT on many similar NLP tasks with less training data [23]. DeBERTa is an encoder-decoder based transformer model [24], based on the architecture of BERT with two key changes. Firstly, disentangled attention handles word content and position vectors separately, rather than adding them together as is the case in BERT. This effectively adds an interaction term from position-to-content in the attention mechanism (see Equation 1). Secondly, the decoder uses absolute and relative word position, whereas BERT uses only absolute position. These changes to the model architecture result in better results on several tasks, even when using half the training text of other LLMs such as RoBERTa [25]. One disadvantage of DeBERTa is that there is no domain-adapted BiooBERT or ClinicalBERT equivalent that has been pre-trained on clinical texts. We tehrefore used the base DeBERTa pre-pretrained model available from the HuggingFace Transformers Hub [26] and its tokenizer as a starting point for further model training, including our own domain adaptation step.

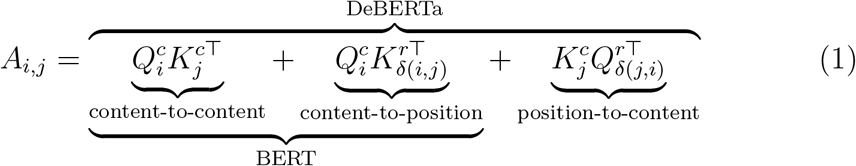

Equation 1: Attention between token i and j is described by the position and content interactions, where 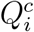 is the content of token i, 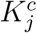 is the content at token j, 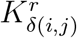 is the relative position of i to j, 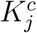 is the content at position j, and 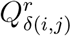 is the relative position of j to i.

#### 2.2.3. Processing and Model Training

Pre-training with clinical texts has demonstrated improvement in other NLP tasks [27], so we therefore performed further pre-training of the DeBERTa model on masked-language modelling (MLM) [17], using a corpus of 1.1 million unannotated clinical documents from our EMR. In this step, 15% of the tokens in the unannotated corpus were masked, with the model trained to predict the mask from the surrounding text. We trained the model for 3 epochs on this task.

Annotated documents were tokenised using a word piece [28] tokeniser and then split into multiple chunks with a maximum token length of 512 tokens. These tokenised documents were then used for fine-tuning the model for the task of NER on drug and ADR annotations (see Figure 2). We annotated ADRs and drugs at the token level for future implementation into our institutional ADR detection workflow, to aid in rapid document assessment by directing attention to the relevant words. However, for algorithm assessment we assessed model performance at the document level. For binary, document-level classification, the token with the highest probability of being an ADR was used. For example, if the word “rash” has the highest probability of being an ADR with a probability of 0.86, then this value is used to classify the document as either containing or not containing an ADR mention. This value is then used for the final model evaluation at the document level using the k-folds cross-validation method.

Model training steps were performed using Python version 3.9.6 [29] and the Transformers library [26] – using a PyTorch backend [30] – on 3 Nvidia 1080ti graphics processing units. Full details of model training, including hyperparameters, can be found in Appendix C. All code is available at https://github.com/AustinMOS/adr-nlp. Experiment tracking is shared at https://wandb.ai/cmcmaster/adr_nlp.

#### 2.2.4. Validation and Testing

Using stratified k-fold cross validation, the annotated discharge summaries were shuffled randomly then split into 5 datasets. Hyperparameter tuning was performed on 1 fold Appendix C. The model was then trained on 4 of the datasets and tested on the 5th (test) dataset. This allowed for the final model to be tested on previously unseen annotated EMR data not used in the training of the model. The process was repeated 5 times to calculate the mean and 95% confidence interval for each metric. The reported metrics are the area under the receiver operating characteristic curve (ROC-AUC) and the area under the precision-recall curve (PR-AUC). A single fold had selected to examine binary classification performance at the Youden’s cutpoint.

In order to validate performance on an unselected cohort, the final model was then evaluated on a randomly selected set of 102 discharge summaries not included in the model development dataset and not enriched for ADRs. These discharge summaries were classified as either containing or not containing an ADR mention, with binary model performance assessed at the Youden’s cutpoint.

#### 2.2.5. Benchmarking

The performance of our model was compared to a previously published machine learning model from our institution [31]. This model was based on a dataset of Y40-59 ICD-10 coded admissions from December 2016 to November 2017. All of these admissions were flagged as possible ADRs and therefore assessed by an expert pharmacist using extensive chart review to determine the veracity of the reports. This model used only ICD-10 coding data and length of stay to discriminate between true and false ADR reports. In addition, the model training procedure was repeated with ClinicalBERT model [27], a large language model that has been pre-trained on several medical data sources, including discharge summaries from the MIMIC III dataset [32]. ClinicalBERT has previously shown excellent performance on other ADE tasks [16] and therefore serves as a strong baseline.

#### 2.2.6. Ablation Study

We also performed an ablation study to test the requirement for the intermediate pre-training step by comparing the final performance of the model with and without this step.

## 3. Results

### 3.1. Data

The pretrained DeBERTa base model was further trained on MLM using a corpus of 1.1 million EMR documents, spanning discharge summaries, progress notes, pathology reports and radiology reports. A further 861 annotated discharge summaries were used for the downstream NER task. All of the annotated discharge summaries came from admissions coded with an ICD-10 Y40.0-Y59.9, with 311 of the 861 containing an ADR mention. The admissions occurred across a broad range of specialities (23% general medicine, 10% geriatrics, 15% surgical).

### 3.2. Model

The model demonstrated good discriminative performance at the document level, with a ROC-AUC 0.955 (95% CI: 0.946 - 0.963) (see Figure 3) and PR-AUC 0.925 (95% CI: 0.902 - 0.948).

**Figure 3:**
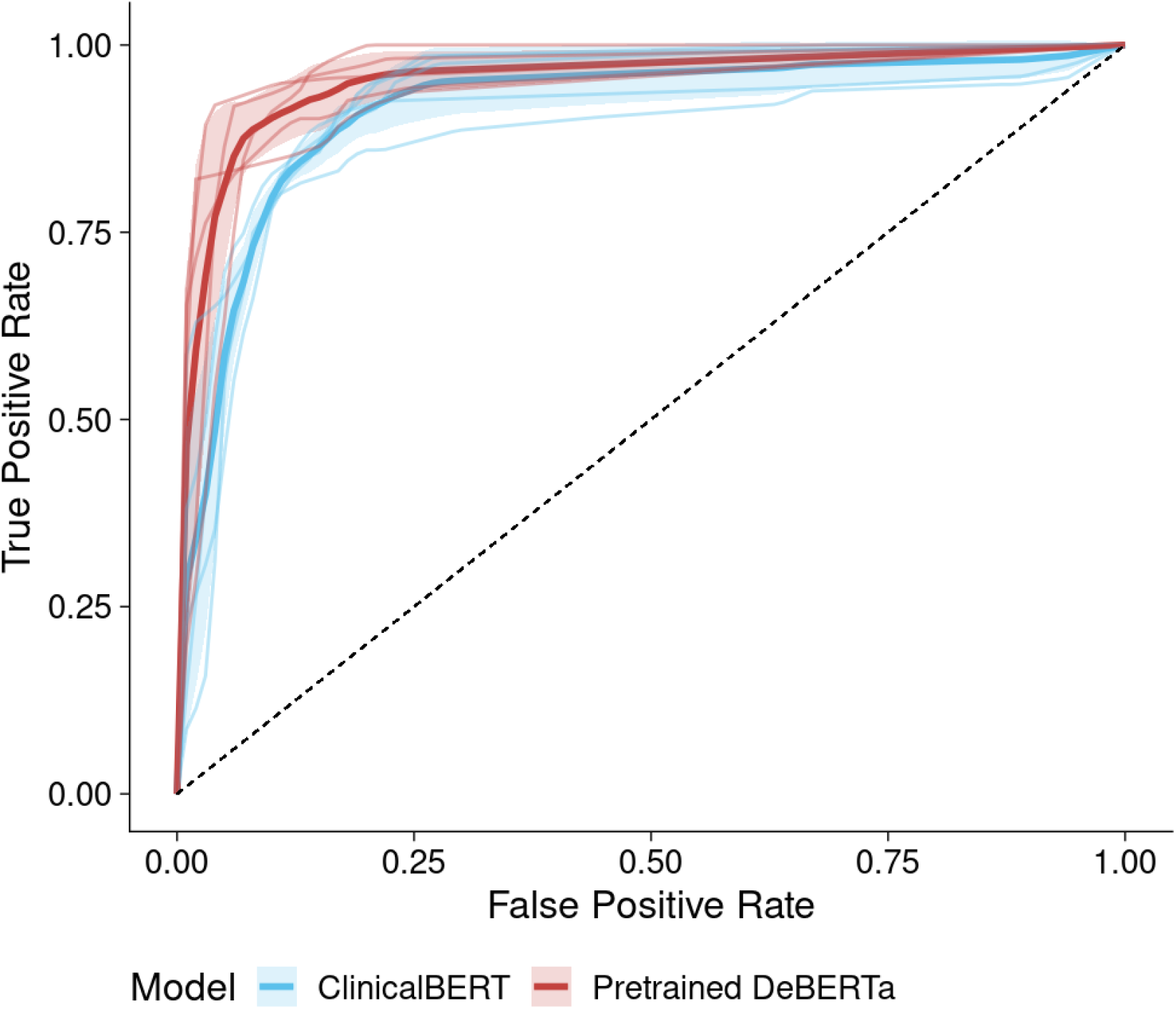
Receiver Operating Curves (Pretrained DeBERTa vs. ClinicalBERT) cross-validation. Mean receiver operating characteristic (ROC) curve of each model is represented by the solid lines, with the shaded areas representing the mean ± standard deviation. The ROC for each individual fold is plotted using thin lines.

There was consistently good performance across all 5 folds with respect to the ROC (Figure 3). The final model was the best, beating both DeBERTa without pre-training and the ClinicalBERT baseline. Intermediate pretraining improved the PR-AUC by 0.31 and the ROC-AUC by 0.34. The final pretrained DeBERTa model had a higher ROC-AUC than the previously published ICD-10 model (see Table 1)).

**Table 1:** Model Performance

| Metric | NLP Model<br>(no pre-training) | NLP Model<br>(with pre-training) | ClinicalBERT | ICD-10 Model |
| --- | --- | --- | --- | --- |
| PR-AUC | 0.894 (0.852 - 0.936) | 0.925 (0.902-0.948) | 0.859 (0.807 - 0.911) | - |
| ROC-AUC | 0.921 (0.884 - 0.958) | 0.955 (0.946-0.963) | 0.917 (0.886 - 0.947) | 0.803 |

The evaluation dataset of 102 unselected discharge summaries had a much lower rate of ADRs (14.7%). Using the Youden J-point threshold for classifying ADRs, the binary classification accuracy was 93.1%, with a sensitivity of 93.3% and specificity of 93.1% (see Figure 4). These results compared favourably to binary classification performance on one of the holdout folds (accuracy 93.1%) 4, although the rate of false positives was higher in the unselected cohort (30%) than the enriched cohort (10.4%).

**Figure 4:**
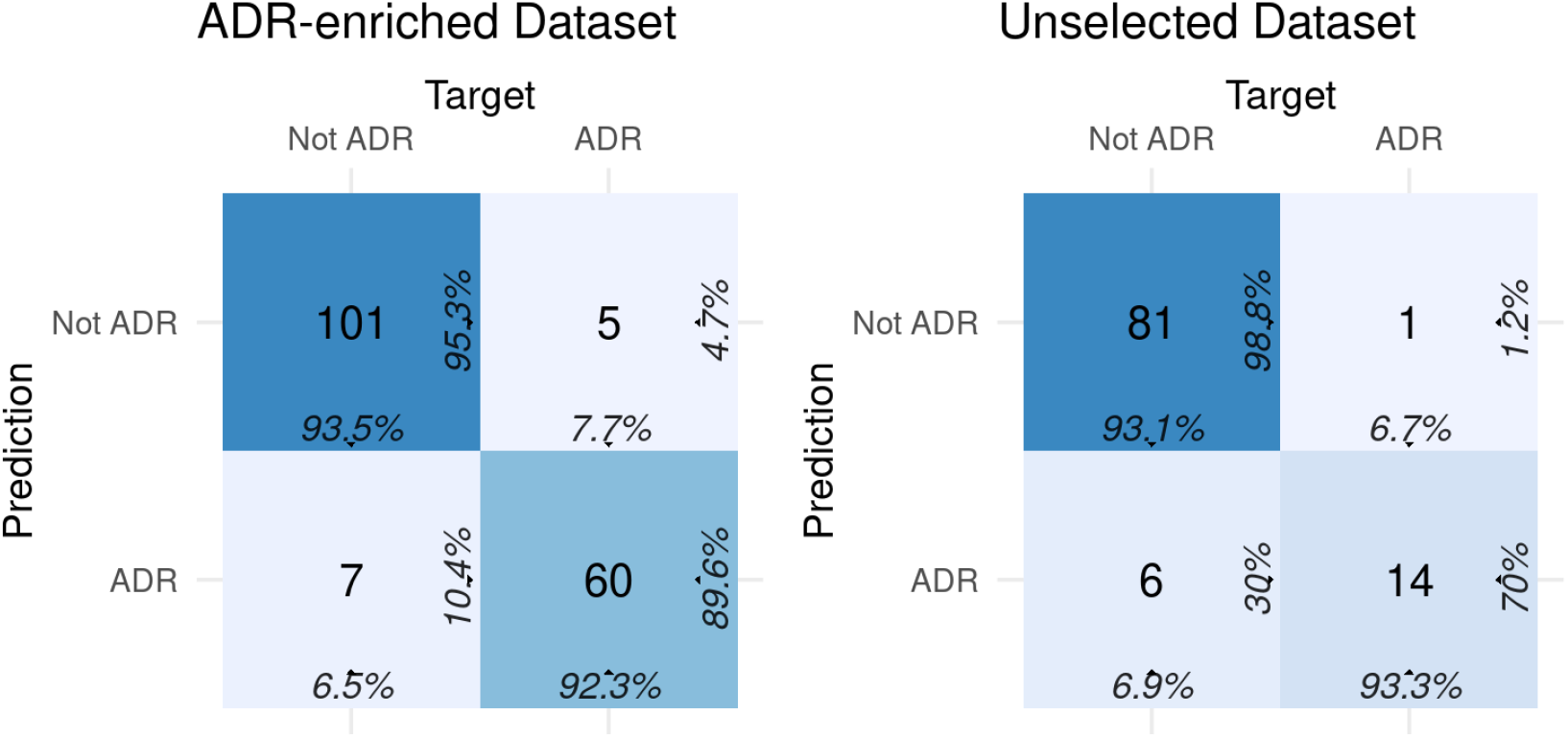
Confusion matrices. Binary performance was similar when the final model was evaluated on a holdout fold from the ADR-enriched dataset and a dataset of unselected discharge summaries. Comparing left and right: sensitivity/recall 0.923 vs. 0.933, specificity 0.953 vs. 0.988, precision 0.896 vs. 0.700, F1 0.909 vs. 0.800, Matthew’s Correlation Coefficient 0.853 vs 0.771.

Examining the 12 misclassified discharge summaries from the holdout fold (see Appendix B) reveals distinct patterns. 5 out of the 7 false positives were adverse drug events, however they all fell short of our annotation criteria because of mild severity not warranting medication cessation. In contrast, 3 out of the 5 false negatives were anticoagulation-related adverse drug events where the anticoagulation was restarted at a lower dose – despite this, these events were annotated as ADRs because of their severity.

Examining the NER output probabilities, the model correctly recognises appropriate entities like “eosinophilia” and “rash” as being common ADR features (see Figure 5). Attribution of these features to a drug increases the ADR class probabilities and this is robust to errors in spelling, even within important words (i.e., drug and ADR feature).

**Figure 5:**
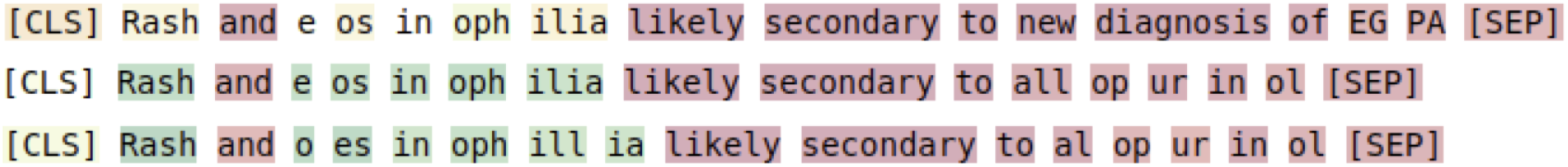
Examples of token-level ADR entity predictions. The color gradient goes from red (very low probability) to green (very high probability), representing the probability that each token belongs to the ADR class.

Exploring modifications to discharge summary text reveals that the algorithm is sensitive to medication mentions (see Table 2). Whilst it appears to be robust to non-Australian medication names like “acetaminophen”, trade names can result in misclassification. This likely reflects documentation practices in our institution, where generic names are much more commonly used. Removing the medication mention, or replacing it with an attribution to a disease or generic term like “a medication” greatly reduces the predicted ADR probability. In comparing these predictions to those produced by the model without intermediate pre-training, the pre-trained model produces robust predictions even when the ADR and causative drug mention are separated, as they may be in dot-point documentation. The model without pre-training requires these entities to be closely related in the text.

**Table 2:** Text-level ADR Predictions. Comparison with and without the additional MLM pre-training step. Output probabilities are scaled from 0 to 1, with cell shading to match (darker shade as probability increases)

| Description | Text | ADR Probability<br>(with pre-training) | ADR Probability<br>(without pre-training) |
| --- | --- | --- | --- |
| Baseline | # Pancreatitis<br>- Lipase: 535 ->154 ->145<br>- Managed with NBM, IV fluids<br>- CT AP and abdo USS: normal<br>- Likely secondary to Azathioprine - ceased, never to be used again.<br>- Resolved with conservative measures | 0.837 | 0.425 |
| Medication line removed | # Pancreatitis<br>- Lipase: 535 ->154 ->145<br>- Managed with NBM, IV fluids<br>- CT AP and abdo USS: normal<br>- Resolved with conservative measures | 0.003 | 0.428 |
| Medication changed | # Pancreatitis<br>- Lipase: 535 ->154 ->145<br>- Managed with NBM, IV fluids<br>- CT AP and abdo USS: normal<br>- Likely secondary to Methotrexate - ceased, never to be used again.<br>- Resolved with conservative measures | 0.997 | 0.424 |
| No specific medication | # Pancreatitis<br>- Lipase: 535 ->154 ->145<br>- Managed with NBM, IV fluids<br>- CT AP and abdo USS: normal<br>- Likely secondary to a medication - ceased, never to be used again.<br>- Resolved with conservative measures | 0.009 | 0.426 |
| Non-Australian medication name | # Pancreatitis<br>- Lipase: 535 ->154 ->145<br>- Managed with NBM, IV fluids<br>- CT AP and abdo USS: normal<br>- Likely secondary to a Acetaminophen - ceased, never to be used again.<br>- Resolved with conservative measures | 0.842 | 0.425 |
| Trade name | # Pancreatitis<br>- Lipase: 535 ->154 ->145<br>- Managed with NBM, IV fluids<br>- CT AP and abdo USS: normal<br>- Likely secondary to a Imuran - ceased, never to be used again.<br>- Resolved with conservative measures | 0.109 | 0.426 |
| Non-medication cause | # Pancreatitis<br>- Lipase: 535 ->154 ->145<br>- Managed with NBM, IV fluids<br>- CT AP and abdo USS: normal<br>- Likely secondary to a alcohol - ceased, never to be used again.<br>- Resolved with conservative measures | 0.015 | 0.426 |
| Single sentence | # Pancreatitis secondary to azathioprine | 0.999 | 0.997 |

## 4. Discussion

We trained a machine learning model on EMR discharge summaries using natural language processing to detect drugs and adverse drug reactions. Compared with a previously published machine learning model [31] using ICD-10 coding data, our study demonstrates an improvement in differentiating true from false ADR reports using NLP. Whilst our model could be deployed to reduce false positives in ICD-10 coding generated ADR reports, by demonstrating good performance on an unselected corpus of discharge summaries, our model could be used across all discharge summaries. This has the benefit of bypassing the intermediate step of generating clinical codes from clinical notes, instead deriving predictions directly from the primary source. This can reduce the inconsistencies, missed ADRs, and human errors which may arise with the use of clinical coding for ADRs. By focusing on clinical text for the identification of ADRs, we return to the fundamentals of ADR detection – the identification of a putatively causal relationship between the administration or ingestion of a drug and a subsequent adverse response, as documented in clinical notes. Although not explored in this study, there is a large potential for future studies to build upon this framework for identifying drugs and ADRs in novel clinical data and train models to recognise temporal relationships between these pairs.

With the increasing transition to recording patient care in electronic formats, machine learning models have the potential to scan large amounts of data for ADRs and improve the accuracy and efficiency of pharmacovigilance within a health network. NLP is an effective method for processing electronic data into structured forms for clinical research. It has been demonstrated to have an accuracy comparable to professional clinical coders in the coding of radiographic reports and is superior to simple text searching methods [33, 34]. The broad applicability of NLP systems is demonstrated by their ability to be extended to recognize new patterns and types of information [35]. The MedLEE NLP system was initially trained for the automated processing of radiological reports, but it has also been successfully extended to detect adverse events in discharge summaries [36, 37]. In a similar vein, our model demonstrates how the open-source DeBERTa model may be extended to develop a machine learning model for ADR prediction. Furthermore, the additional step of pre-training on a larger corpus of discharge summaries from our institutional EMR lead to improved model accuracy. Our model outperformed a finetuned model based on ClinicalBERT, a language model that has been pre-trained on a large corpus of discharge summaries and has demonstrated state-of-the-art performance in other ADE tasks. We hypothesise that this is because our model has learnt some of the linguistic features that are unique to our regional (Australia) and local (institution) context.

In recent years there has been increasing interest in using NLP models for the detection of ADEs. The open NLP challenge for ADE detection was held in 2018, using the MADE 1.0 corpus of annotated clinical notes related to drug safety surveillance [27]. However, these datasets have not been developed specifically to identify ADRs and distinguish them from other ADEs. This is especially important given the high rates of ADR under-reporting and their relatively rare occurrence in clinical notes. Therefore, a major strength of our model was the use of real-world clinical ADR reports in the annotation of our dataset. Our model was trained on 861 Y40.0-Y59.9 ICD-coded discharge summaries, of which 311 were labelled as ADR reports, with a significant subset (231 admissions) having been validated as true ADRs by our institutional ADR committee. This resulted in a smaller class imbalance than would have been observed had we used an unenriched sample of discharge summaries.

A key potential limitation of any NLP algorithm is the quality of the training data provided. For instance, a model will perform poorly where there are inconsistent annotations. A labelling scheme in our study was therefore used to reduce ambiguity and guide the manual annotation process for the training of our model. Although all annotations were reviewed by a senior clinician, the inherent biases of a single annotator are also carried forward into the biases of the final model and its prediction of ADR and drug labels. The misclassified labels we examined did demonstrate inconsistencies in labelling, where 8 of 12 misclassified discharge summaries were cases in which the offending medication was restarted – some labelled as ADRs and some not. Whilst these are not ADRs for the purposes of a patient drug allergy history, when they fall on the severe end of the spectrum they are important to recognise for the purpose of population-level pharmacovigilance. More prescriptive annotation criteria might overcome this problem, however it might also mean that important severe events are missed. One way to overcome this might be by making event severity its own annotation and combining ADR and severity predictions to ensure severe events with less certainty about causality are captured for manual review.

In contrast to other work that has used public ADE datasets, we created our own dataset in order to develop an algorithm that had validity within our hospital and could therefore be deployed in clinical practice with confidence. Annotating such a datset for NER tasks can be time-consuming and costly. The ability to quickly and accurately annotate a corpus of this size was accelerated using active learning, allowing full review of each document whilst reducing the cost of producing further annotations with each subsequent batch. Although we did not reach a point in which we had saturated model improvement (4% improvement in token-level accuracy when using 100% vs. 75% of the corpus), increasing the corpus size beyond 861 is unlikely to result in significant improvements alone.

In terms of our dataset, our model was trained solely on ICD-10 coded Y40-Y59 discharge summaries rather than the full spectrum of EMR data. In order to be included in the training of the model, ICD-10 coding relies on the clinicians and clinical coders to have documented and identified an event that was thought to be an ADE. Although this may have missed some important confounding features in unselected discharge summaries, we are reassured by the consistent performance in the unselected evaluation dataset. Beyond discharge summaries, expanding the dataset to encompass other EMR notes (e.g. inpatient notes, pathology reports etc.) is another direction for future study. Additional improvements may be made to this model by annotating and training the model to identify linked concepts, such as drug indication and dose, separate annotations for non-ADR ADEs, and further training of the model to recognize drug-ADR and drug-dose-indication relations.

Whilst there is often consistency in the formatting of discharge summaries between health networks, it will require validation on these datasets to demonstrate generalizability. In particular, it is likely that we will observe a decrease in performance in institutions where trade name documentation is more prevalent. We anticipate that this problem can be prevented with synthetic training data generated from our current dataset, by simply replacing generic names with trade names from a thesaurus of generic-trade name pairs. Although it is beyond the scope of this work, we plan to perform external validation on data from other hospitals to explore external validity and identify differences in documentation that may limit this.

## 5. Conclusion

Our study demonstrates the potential for NLP models to be developed for automated ADR detection in a real world setting. This approach can address the under-reporting issues of current methods, bypass the resource limitations of current clinical workflows and increase the ADR reporting rates within the hospital. With additional pre-training on EMR data specific to our health network, the model was able to learn the patterns of discharge summary formatting, allowing correct classification even of distant relations when documented within the expected structure of a discharge summary. The unique construct of our corpus, particularly the presence of many validated ADRs alongside non-ADR ADEs, meant that our model had to differentiate ADRs from other incidents of medication-related harm. We plan to implement our model into the clinical workflow of ADR reporting. Specifically, we are working on an ADR dashboard to present ADR reports, derived from both spontaneous reporting and our NLP model. Reports will be presented with the relevant section of the discharge summary highlighted according to the NER outputs of the model. These annotations can be confirmed or rejected by the pharmacist reviewing the report, providing ongoing annotations to further train and refine our model. We hope to identify any dataset drift by this method, including noval ADRs, new medications and new documentation practices.

## Data Availability

Data are not available for external requests

### Appendix A: NER Annotation Label Scheme

1. Only label drugs that are possibly responsible for an ADR
2. ‘Drug’ labels are only given for drugs administered at therapeutic dose.
3. ‘Drug’ labels can be given if it is a clear cause of an ADR even if it is unclear what ADR occurred.
4. Preference labelling of specific drugs. Drug class can be labelled if specific causative drug is not mentioned (e.g. “angioedema after starting ACE-inhibitor”)
5. ADR labels are only assigned to disease states, signs and symptoms, not pathological/radiological findings with no clinical consequence.
6. Abbreviations for drugs and ADRs are permitted
7. Preference drug and ADR labels which are in close proximity within the document
8. Preference labels with causal language – e.g. ‘due to’, ‘secondary to’, ‘withheld’, ‘ceased’, ‘complicated by’.
9. Multiple ADRs may be labelled for a given drug
10. Multiple drugs may be labelled as (potentially) responsible for a given ADR
11. Do not label drugs and ADRs already recorded in the patient allergy list
12. Only label drugs documented in the body of the text, not in a drug list

### Appendix B: Model Errors

#### False Negatives

- 1 instance of interstitial nephritis lacking drug attribution in the body of the text (likely association with NSAIDs only mentioned on the discharge plan at the very end of the document)
- 3 instances of anticoagulation being restarted after a bleed – all could conceivably be considered adverse drug events and not ADRs
- 1 instance of transaminitis from statin (only instance of statins being implicated in dataset, only instance of transaminitis)
  - Transaminitis still assigned probability of 41

#### False Positives

- Elevated transaminases in the setting of therapy for Mycobacterium tuberculosis
  - Could reasonably have been labelled as an ADR, but not done so because self-resolved without any changes to therapy
- Lithium toxicity already recognised prior to admission with plan to restart at a later date
  - Given plan to restart, not an ADR that could be placed on the patient’s record, but important to note
- Past history of chemotherapy-induced pancytopenia, not an ADR relevant to current admission
- Multifactorial anaemia - partially attributed to anticoagulation (not ceased)
- Headache requiring lumbar puncture - anticoagulation withheld for the procedure
- Peripheral oedema secondary to amlodipine (not ceased) - adverse drug event that could conceivably be labelled as an ADR
- Possible medication-induced bradycardia vs. sick sinus syndrome - decision made to continue beta-blocker and tolerate slower rate – likely medication-induced, but not rising to the severity of an ADR

### Appendix C: Model Training Details

#### Pre-training

The following hyperparameters were used:

- Learning rate: 5e-5
- Weight decay: 0.01
- Training duration: 3 epochs (12 hours)

#### Fine-tuning

Hyperparameter tuning:

- Learning rate: {1*e*^*−*6^, 5*e*^*−*6^, 9*e*^*−*6^, 1*e*^*−*5^, 5*e*^*−*5^}
- Training epochs: {5, 10, 15, 20, 25}
- Gradient accumulation: {1, 2, 3, 4, 5}

All final hyperparameters can be found in the configuration parameters stored with each experiment at https://wandb.ai/cmcmaster/adr_nlp.

#### Pre-trained Models

The pre-trained models used in this work are availabel from the following links:

- DeBERTa Base: https://huggingface.co/microsoft/deberta-base
- ClinicalBERT: https://huggingface.co/emilyalsentzer/Bio_ClinicalBERT

